# Superspreading as a Regular Factor of the COVID-19 Pandemic: I. A Two-Component Model

**DOI:** 10.1101/2020.06.29.20138008

**Authors:** Juri Dimaschko

## Abstract

We consider the impact of superspreading on the course of the COVID-19 epidemic. A two-component model of the epidemic has been developed, in which all infected are divided in two groups. The groups are asymptomatic *superspreaders* spreading the infection and *sensitive* persons which can only get infection. Once infected the *sensitive* exhibit clear symptoms and become isolated. It is shown that the ratio of increment of the number of daily cases in the beginning of the epidemic and decrement at the end of the epidemic is equal to the ratio of the spreading rates of the infection transmission from the *superspreaders* to potential *superspreaders* and to the *sensitive* persons, respectively. On the basis of data from 12 countries and territories it is found that the *superspreaders* transmit the infection to potential *superspreaders* approximately 4 times more often then to the *sensitive* persons. Specific measures to limit the epidemical incidence are proposed. The possibility of an allergic component in the disease is discussed.

## I. INTRODUCTION

The COVID-19 pandemic has a number of differences from previous flu epidemics.

1. One of its main features is the existence of active asymptomatic carriers of the virus - *superspreaders* [1]. They cause a significant contribution to the spread of the virus because once infected do not sick and therefore are not isolated. Such carriers can be detected by random testing or by testing of contacts of already infected persons (which, of course, is more effective).

2. Another feature of the COVID-19 pandemic is the significantly lower number of cases (about 0.2 – 2%) than the normal flu epidemic (10 – 20%). This suggests that only a small fraction of people are sensitive to the SARS CoV-2 virus that is responsible for the current pandemic. As in the case of the *superspreaders*, they have also been successful in reproducing the virus, but this is now accompanied by a noticeable immune response and thus a disease.

The aim of this work is to take into account the impact on the spread of the COVID-19 epidemic of both factors.

1. The presence of a limited number of people who are susceptible and insensitive to the virus and who, once infected, become asymptomatic carriers and spreaders of the virus;

2. The presence of a limited number of people who are both susceptible to and sensitive to the virus and who, been infected, exhibit a strong immune response and get sick.

This approach treats *superspreaders* not as an exotic factor leading to separate outbreaks of the virus, but as a regular element in the dynamics of the epidemic. It is this element that defines the main features of the development of the COVID-19 pandemic.

The article is structured as follows.

The second part classifies those infected by the nature and intensity of the immune response, which is the basis for the further proposed epidemiological model.

In the third section, a corresponding two-component model of the epidemic is constructed and its analytical solution is found.

The fourth part compares the found results with observed epidemics in a number of countries and territories.

The final part summarizes the application of the model and discusses the possibilities for limiting the incidence resulting from this model.

## II. IMMUNOLOGICAL CLASSIFICATION OF INFECTED

From an epidemic point of view, the mobility of infected persons is the most important factor, as only the mobile infected persons can transmit the infection. It is therefore reasonable to divide those infected into two broad groups by degree of mobility. Since mobility, in turn, is determined by the presence of pronounced symptoms of disease that are linked to immune response, such separation must be based precisely on the nature and degree of the immune response.

During the COVID-19 pandemic, a significant amount of data was collected describing the nature and diversity of the human immune response to the virus. In this section, we will use part of this data to limit and give an immunologically accurate definition of the two main groups of the model - *superspreaders* and *sensitive*.

The basis of the definition is the appearance of antibodies to the virus in the mucous membrane (IgA immunoglobulin), as well as in the blood (IgG immunoglobulin). Tests taken in persons with different degrees of severity, from asymptomatic to acute, show the following pattern [2].

1. For asymptomatically infected persons, both types of antibodies are largely absent. This fact points to the category of *superspreaders* and indirectly explains the large number of viruses in such carriers, resulting in numerous cases under certain circumstances.

2. For those infected in mild form, antibodies are found mainly in the mucosa (immunoglobulin IgA). These infected individuals must be classified as *sensitive*, as the presence of signs of a disease confirmed by a positive virus test results in their immediate isolation.

3. For those who have suffered an acute infection, antibodies are found both in the mucosa and in the blood (IgA and IgG immunoglobulins). These infected people should also be classified as *sensitive*, as they are usually hospitalized and anyway isolated. Unlike those who have experienced the disease in mild form, we will further refer this part of the category *sensitive* to the subcategory *supersensitive*. This separation does not affect the proposed two-component model, which is concerned only with the fact that the carrier is isolated.

In this way, one can give an immunologically precise definition of the groups *superspreaders* and *sensitive*. The first includes only asymptomatic carriers, which are not subject to any isolation and spread the infection. The second is symptomatic, isolated and therefore unable to spread the infection. This division is presented in the Table 1. The *sensitive* group has a subgroup *supersensitive*. It includes those infected who exhibit the strongest immune response, including antibodies in both the mucous membrane and the blood.

**Table 1:** Definition of two groups (*superspreaders* and *sensitive/supersensitive*) by the type of immune reaction

|  | Virus | IgA | IgG | Mobility |
| --- | --- | --- | --- | --- |
| *superspreader* | + | - | - | free |
| *sensitive* | + | + | - | isolated |
| *supersensitive* | + | + | + | isolated |

From an epidemic perspective, *sensitive* and *supersensitive* groups are equivalent, as in both cases the infected persons are isolated and unable to spread the infection further.

## III. THE TWO-COMPONENT MODEL OF THE EPIDEMIC

Consider an epidemic model involving only two types of potential carriers. These are *superspreaders* and *sensitive* persons. The *superspreader*, once infected, remains asymptomatic. He does not get sick and begins to spread the infection. *Sensitive* persons, been infected get sick, isolated, and unable to transmit the infection. Thereby in accordance with classification in Table 1, the *super-sensitive* persons in this model are included in the group of *sensitive*.

Unlike the conventional one-component model of the epidemic, where anyone infected continues the chain of infection, in this model only *superspreaders* have this capability. The *sensitive* persons play a passive role only.

They can get the infection but not spread it. Thus, the epidemic is two-component. The two components of the epidemic, *sensitive* and *superspreaders*, are progressing in parallel. The latter has a unilateral impact on the former, while the former cannot influence the latter.

Let the full number of the *sensitive* persons be *N*_1_, of which *n*_1_ are infected by time t. The total number of the *superspreaders* is *N*_2_, of which *n*_2_ are infected by time t. Then the dynamics of the epidemic is described by a system of equations

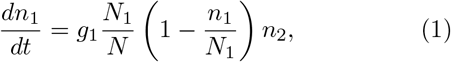

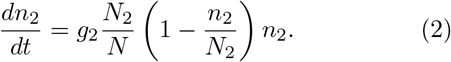

where *N* is the total population, and *g*_1_*,g*_2_ are spread rates. They are average numbers of *sensitive* and potential *superspreaders* which can be infected by one *superspreader* in one day.

The definitions of all variables and parameters of the model are given in Table 2, and the scheme of interaction between the groups of the *superspreaders* and *sensitive* leading to the dynamic system of equations (1, 2), is presented in Fig.1.

**Table 2:** Definitions of model variables and parameters

| Variables and parameters | Definition |
| --- | --- |
| $n_1$ | Current number of infected *sensitive* persons |
| $n_2$ | Current number of infected *superspreaders* |
| $N_1$ | The total number of *sensitive* persons |
| $N_2$ | The total number of *superspreaders* |
| $t$ | Time (in days) |
| $g_1$ | Spread rate to *sensitive* |
| $g_2$ | Spread rate to potential *superspreaders* |

**Fig. 1:**
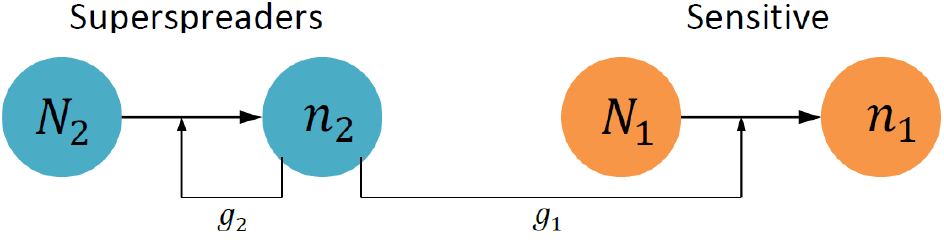
Scheme of the two-component epidemic model

After introducing new dimensionless variables

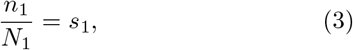

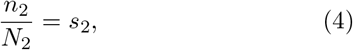

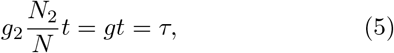

the initial system of equations (1,2) acquires a form of

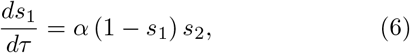

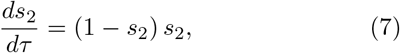

where

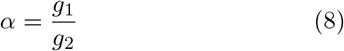

is a *Relative Spread Rate (RSR)*, and parameter *g* = *g*_2_*N*_2_/*N* introduced by (5) is an effective spread rate.

The resulting system of equations (6, 7) has an exact solution

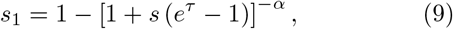

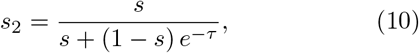

that meets the initial conditions

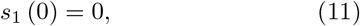

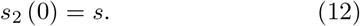

The given solution corresponds to the initial state, in which there is already a non-zero number of infected *superspreaders*, *n*_2_ = *sN*_2_, but no sick persons yet,*n*_1_ = 0. The empirical value observed is the daily number of cases *dn*_1_=*dt*, which according to the solution (9) depends on time as follows (see Appendix A):

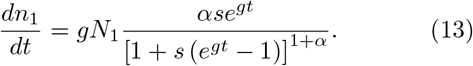

An important property of this solution is the asymmetry of its exponential asymptotic at the beginning and end of the epidemic, i.e. for *t* → 0 and for *t* → 1. At the beginning of the epidemic, it behaves as

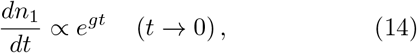

and at the end of the epidemic as

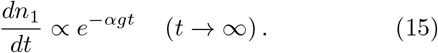

Three graphs in Fig.2 demonstrate the growth of this asymmetry as *α* parameter decreases. The smaller the value of the parameter *α*, the slower the daily number of cases decreases. This is quite natural, because the increase of *dn*_1_/*dt* is due to activation of the *superspreaders* with the rate of *g*, whereas the decline is due to exhausting the fraction of susceptible among the *sensitive* with the rate of *αg*.

**Fig. 2:**
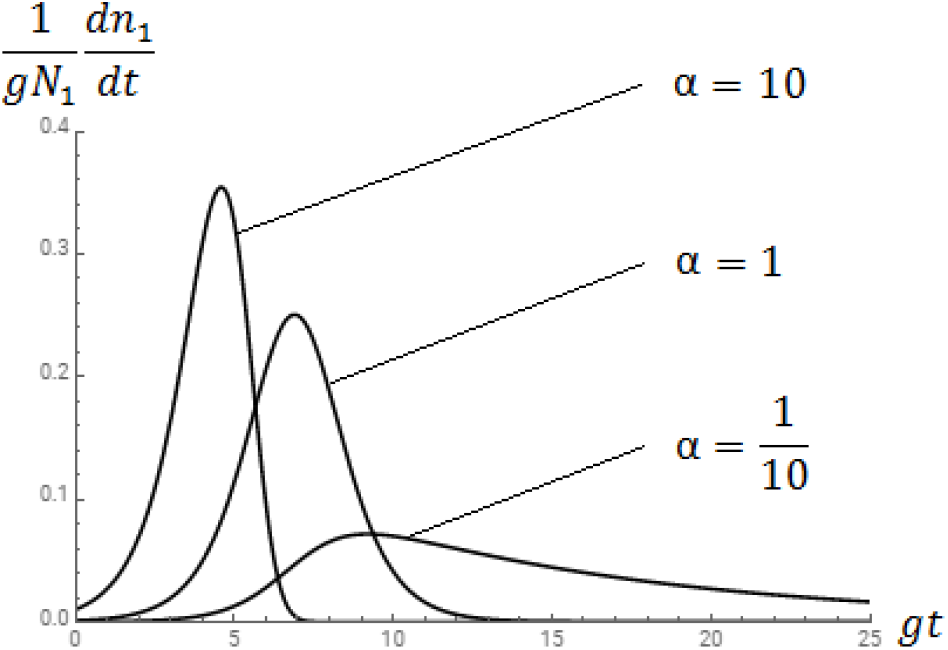
Dependence of the daily number of cases on time at different values of *α* = *g*_1_=*g*_2_

As follows from the asymptotic (14, 15), the ratio of the exponential decrement at the end of the epidemic and the increment at its beginning directly gives the parameter *α* = *g*_1_/*g*_2_, i.e. the *Relative Spread Rate*.

The analytical solution (9, 10) enables to find the exact position of the maximum point for the daily number of cases, as is done in Appendix A. If the spread rate constant *g* increases, the maximum is reached earlier. The increase in the number of *superspreaders*, expressed in the growth of the parameter *α* = *g*_1_/*g*_2_, has the same effect, as shown in Fig.2.

The found solution (13) is invariant under following rescaling of parameters and variables: *g*_1_ *→ g*_1_λ*,g*_2_ *→ g*_2_*λ*, *N*_2_ *→ N*_2_*/λ,n*_2_ *→ n*_2_/*λ*, because the parameters of this solution *(g, N*_1_*, a* and *s)* remain unchanged. Thereby (13) gives no concrete value of the number of *superspreaders*. On the other hand, this solution enables to reproduce the observable dependence of the daily number of cases on time and to find (by fitting) all four its parameters.

Comparing the course of two epidemics, the Spanish flu, in 1918. and the current COVID-19 epidemic, shown in Fig.3, exhibits a marked difference between them in the final phase of the decline. In the case of COVID-19, it occurs much more slowly. The two-component epidemic model can explain this by assuming a relatively small *Relative Spread Rate, α* ≪ 1, as shown by the third graph in Fig.2.

**Fig. 3:**
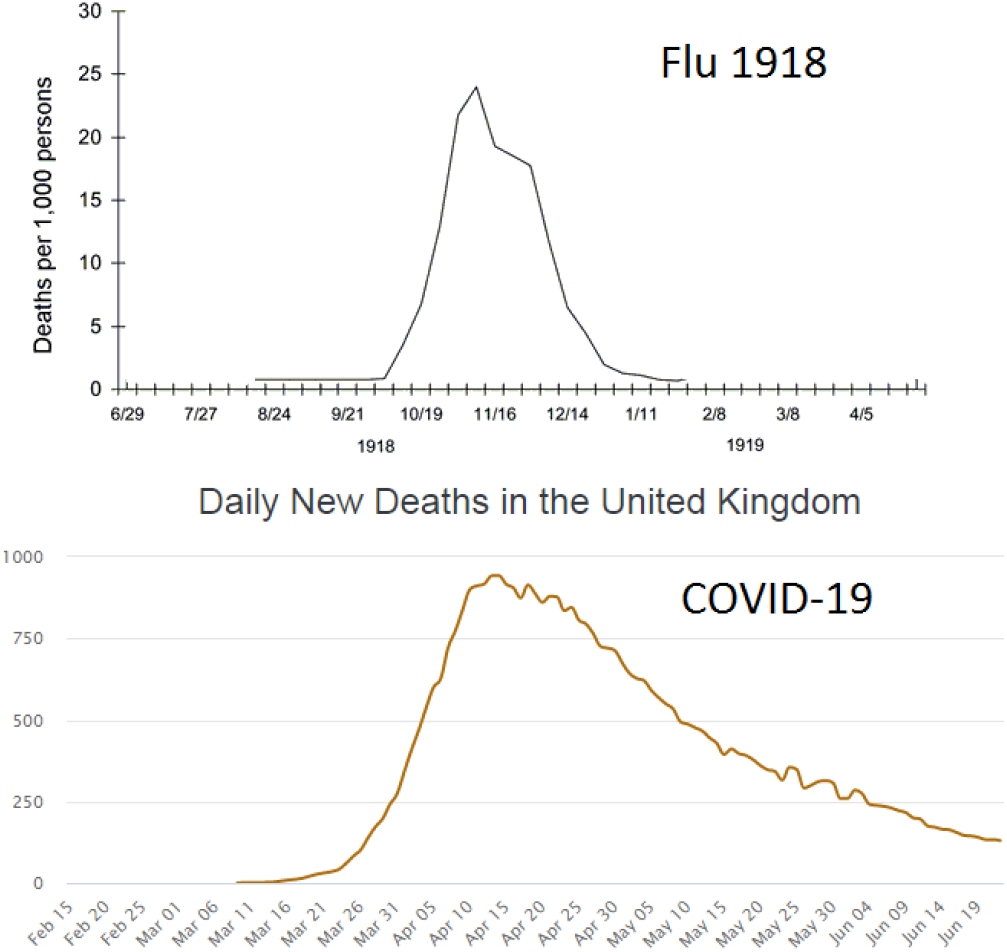
Course of two epidemics in the UK: u (1918) and COVID-19 (2020)

In the proposed model, the virus first undergoes exponentially rapid spread among potential *superspreaders* and sequential activation, which means infection, most of them. Then activated *superspreaders* continue to spread the virus among the *sensitive* persons. The number of spreaders at the point of maximum is already saturated, and the base for further spreading of the virus among *sensitive* is also gradually being exhausted. Thereby the epidemic is subsiding.

## IV. ANALYSIS OF THE EPIDEMIC IN DIFFERENT COUNTRIES AND STATES

First of all, in many countries, as in the world as a whole, the COVID-19 epidemic is accompanied by a crossover of the very type of dependence of the number of infected on time. For a short time, about two weeks, it changes from exponential to purely linear growth. One possibility to explain this phenomenon was presented in [3], whose authors connected it with a phase transition from the mean field mode to the mode of personal interaction, when the number of contacts for each carrier becomes less than some critical value.

However, the two-component model can provide a simple and transparent explanation of the observed exponential-to-linear crossover. Indeed, if every infected person can transmit an infection to anyone, then the increase in the number of infected can be only exponential. The first 10 infected people infect 20, the latter 40, then 80, and so on in geometric progression until the base of susceptible is exhausted.

On the other hand, if only a relatively small fraction of all susceptible people, *superspreaders*, can spread the infection, then they are first exponentially activated. After that, the number of active *superspreaders n*_2_ reaches the limit of *N*_2_ - for example, 1000 persons - and no longer grows. Now every day they infect the same number of people, and therefore the total number of infected grows over time, not in geometric but in arithmetic progression - 1000, 2000, 3000, etc. This linear law persists until it is affected by a reduction in the base of people who are not yet infected and sensitive to the infection.

The only condition for the appearance of the extended linear segment in the *n*1(*t*) dependence is a relatively small value of the *Relative Spread Rate*, *α* ≪ 1. It is this case that is represented by the third graph in Fig.2. Next, consider the course of the epidemic in a number of countries and states where it has already peaked and is expected to end soon. In each case, we fit the empirical daily number of cases by the result (13) of the two-component model.

Fig.5 shows the results of this fitting for 12 countries and states: Austria, Belgium, Canada, France, Germany, Italy, Netherlands, New Jersey and New York, Spain, Switzerland and the United Kingdom. The empirical values are shown by points, and the model result is shown by a red line. In each case both the Relative Spread Rate *α* = *g*_1_=*g*_2_ and the effective spread rate *g* = *g*_2_*N*_2_ = *N* were found. The parameters following from this fitting are summarized in Table 3.

**Table 3:** Model parameters for some countries and states. Here N is the population of the country or state, *N*_1_ is the total number of *sensitive*, 100% × *N*_1_/*N* is the asymptotic incidence, *α* = *g*_1_ = *g*_2_ is the *Relative Spread Rate*, *g* = *g*_2_*N*_2_ = *N* is the effective spread rate.

| Country/State | $N$ | $N_1$ | $100\% \cdot N_1/N$ | $\alpha = g_1/g_2$ | $g$ (1/d) |
| --- | --- | --- | --- | --- | --- |
| Austria | 9.000.000 | 16.000 | 0,18% | 0,31 | 0,29 |
| Belgium | 9.000.000 | 60.000 | 0,67% | 0,32 | 0,16 |
| Canada | 38.000.000 | 120.000 | 0,32% | 0,29 | 0,10 |
| France | 65.000.000 | 155.000 | 0,24% | 0,23 | 0,21 |
| Germany | 84.000.000 | 185.000 | 0,22% | 0,24 | 0,24 |
| Italy | 60.000.000 | 250.000 | 0,42% | 0,20 | 0,20 |
| Netherlands | 17.000.000 | 50.000 | 0,29% | 0,32 | 0,15 |
| New Jersey | 9.000.000 | 185.000 | 2,06% | 0,19 | 0,18 |
| New York | 19.000.000 | 420.000 | 2,21% | 0,21 | 0,22 |
| Spain | 47.000.000 | 300.000 | 0,64% | 0,13 | 0,27 |
| Switzerland | 9.000.000 | 30.000 | 0,33% | 0,30 | 0,25 |
| UK | 68.000.000 | 350.000 | 0,51% | 0,19 | 0,14 |

In all cases shown in Fig.5, the exponential growth of daily morbidity at the start of the epidemic is much faster than its further decline. The reason for this is that in all cases the *Relative Spread Rate α* = *N*_2_=*N*_1_ is less than one. The very narrow range of values of the parameter *α*, mainly from 0.2 to 0.3, is noteworthy, while the incidence varies much more widely - from 0:2% to 2%.

**Fig. 4:**
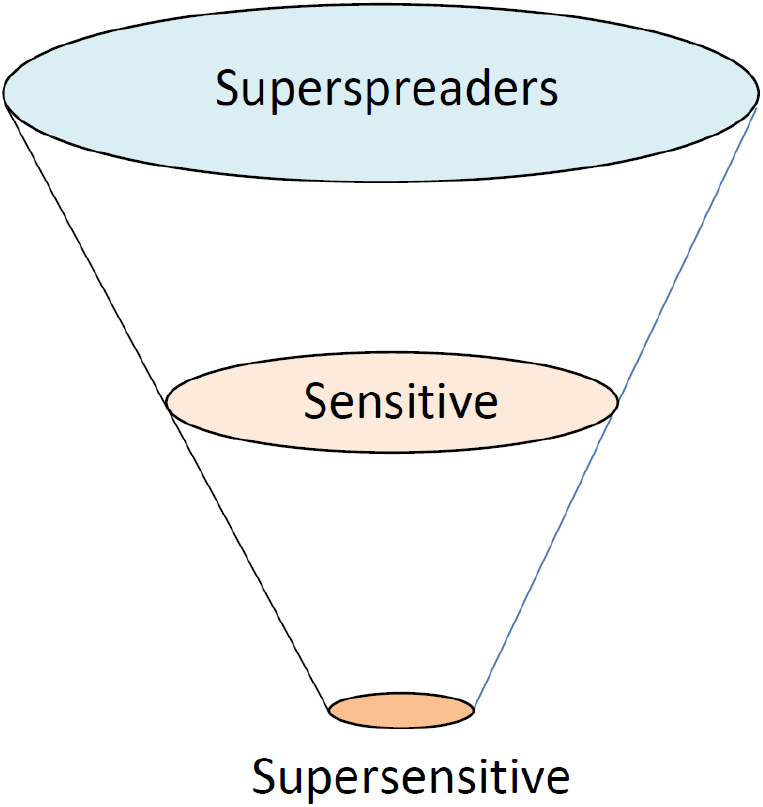
Possible hierarchy of COVID-19 according to the strength of the immune response

**Fig. 5:**
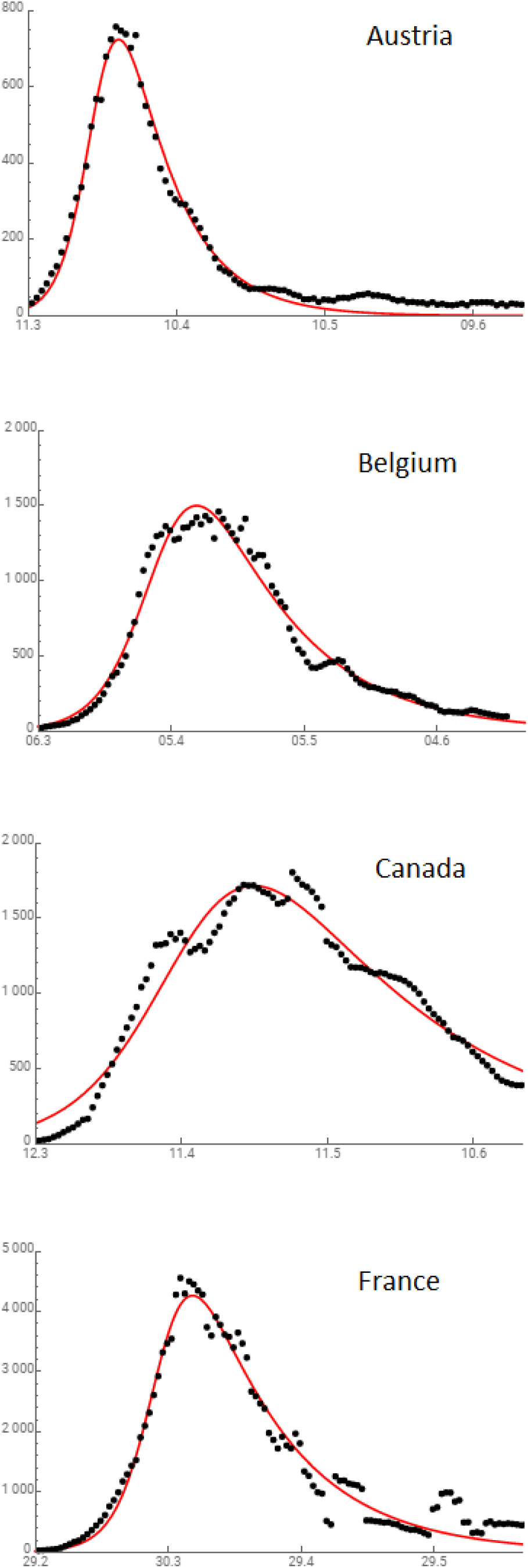
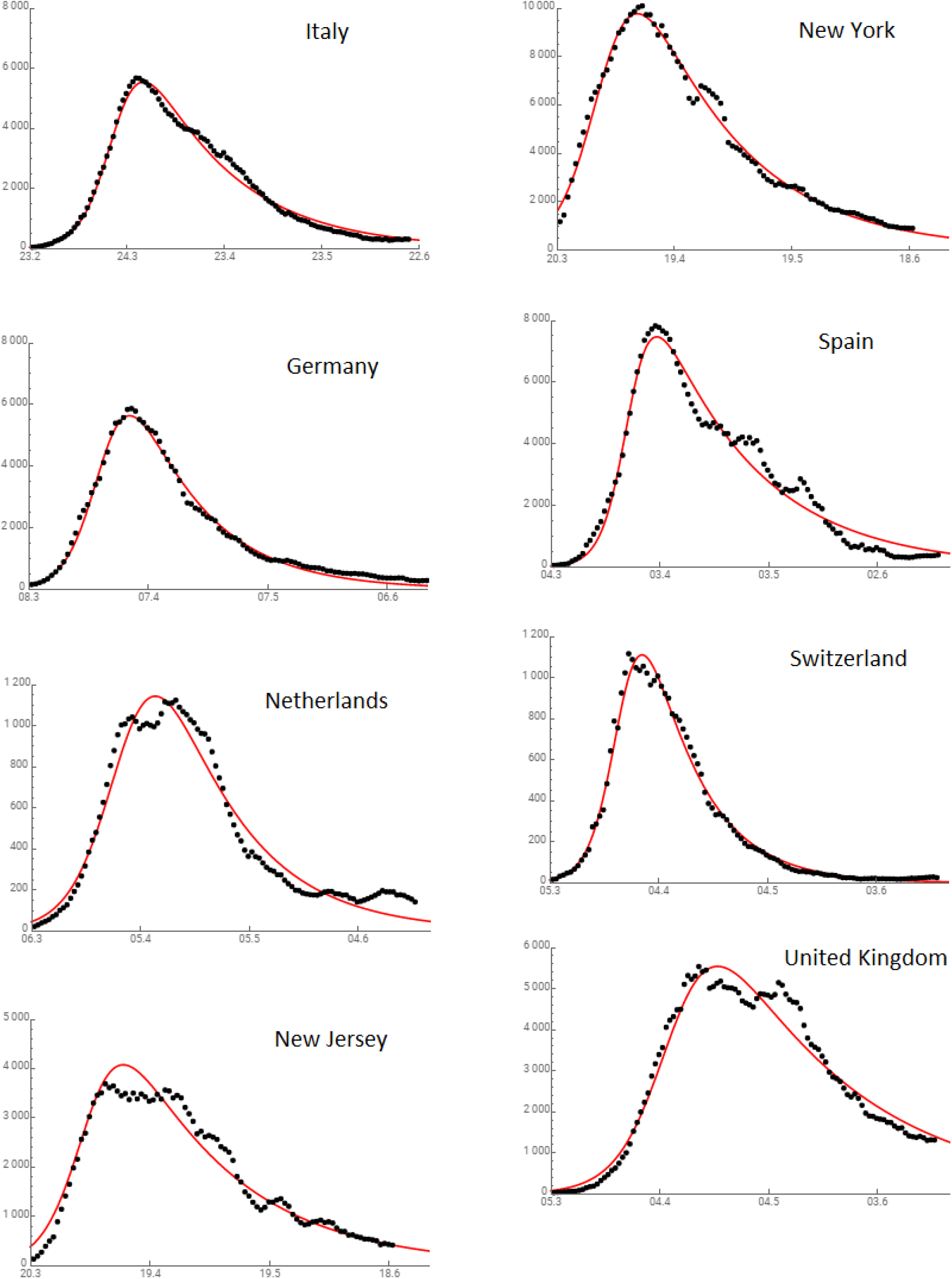
Comparison of course of COVID-19 epidemic in 12 countries and territories (points) with two-component model (red line)

Based on the found tipical values of the *Relative Spread Rate α* = 0.24±0.06 one can conclude, that *superspreaders* transmit the infection to potential *superspreaders* approx. 4 time more often then to the *sensitive* or *supersensitive*. Apparently this means, that absolute majority of infected are asymptomatic *superspreaders* exhibiting no clear immune response. On the other hand, the infected *supersensitive* are absolute minority among all infected.

Following to the initial classification, see Fig.1, one can suggest a hierarchy of disease according to the strength of the immune response shown in Fig.4.

## V. CONCLUSIONS AND DISCUSSION

Thus, despite all its simplicity and schematicity, the proposed two-component model gives a plausible description of the course of the COVID-19 epidemic in all considered cases.

The proposed model is based on two specific types of virus carriers. They are

1. *superspreaders*, in which the infection results in active reproduction of viruses without appreciable immune response and any symptoms of the disease, and
2. *sensitive* persons, in which infection leads to the reproduction of viruses, and to a noticeable, and in the case of *supersensitive* often to an acute immune response.

It is this reaction that manifests itself as the terminal course of the disease.

The rest of the population is neutral in relation to the virus - in most cases the virus appears not to be reproducible in significant amounts in cells. It should be noted that this is the absolute majority, currently 99:84% of the total population of the Earth.

The comparison of possible variants of the body’s immune response to the COVID-19 pathogen presented in the Table 1 raises the question of which of these variants is adequate. There are two possible answers to this question:

1. The immune response is generally adequate to the degree of damage that the virus causes to the cell. In the case of *superspreaders*, the virus for some reason does not cause significant damage to the cells, and therefore there is no immune response. In the case of *supersensitive*, the virus causes significant damage, which causes a strong immune response and leads to the terminal course of the disease.
2. The immune response is largely inadequate to the degree of damage that the virus causes or may cause to the cell. In the case of *superspreaders*, the immune system ignores the appearance and reproduction of the virus and the person remains virtually healthy. In the case of *supersensitive*, a strong immune response occurs and the person is exposed to the disease in an acute form.

Both approaches describe the same infection with the same result, the difference between them is in the interpretation of the nature of the immune response. In the first case, it is considered as an adequate response to the virus. In the second case, the immune response is interpreted as an inadequate allergic reaction to the very presence of the virus, or to the products of its activity. And then the allergic component plays a significant role in the course of the disease.

In the first case, a viral infection is the cause and the immune response is a consequence of the disease. In the second case, the viral infection is a virus agent that provokes an inadequate response of the immune system.

The answer to this question determines what measures can reduce the incidence.

In the first case, it is necessary **to activate** a specific immune response through a proper vaccination. The target group of the vaccination here is *superspreaders*. Then the reduction of incidence is achieved by reduction of the number of the *superspreaders* via transferring them to an intermediate group of *sensitive*.

In the second case, the aim of a proper vaccination is **to reduce** a specific immune response of the *supersensitive*. Then the target group of the vaccination is *supersensitive*, and the reduction of incidence and lethality is achieved by transferring the *supersensitive* to the more safe intermediate group of *sensitive*.

And in both cases it seems reasonable to take preventive measures to early identify *superspreaders* and *supersensitive*, followed by limiting their contacts.

Extending the two-component model by taking into account the deactivation of *superspreaders* over time does not lead to any qualitative change in the results. However, this deactivation turns out to be important in the transition processes after the abolition of quarantine and in the emergence of the second wave. These issues will be discussed in the next article.

## Data Availability

All data are available.

## ACKNOWLEDGEMENTS

I am deeply indebted to Prof. Mykola Iabluchanskiy and Vadym Mogilevsky for valuable remarks and discussions, and to Dr. Daniel Genin for help in use of Mathematika tools.

## Appendix A

As follows from formula (10) of the main text for *s*_1_ ist derivative is

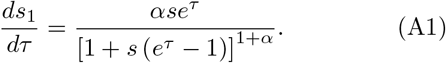

After replacing the variables (3-5), this gives formula (13) for the daily number of cases.

To find the point of the maximum, we write out the logarithm of *ds*_1_=*dτ* and turn to zero its derivative:

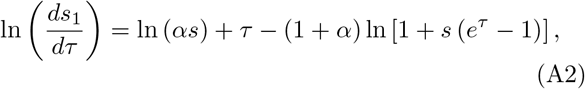

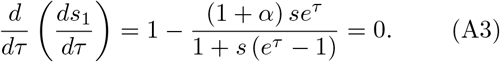

This directly follows the point of maximum:

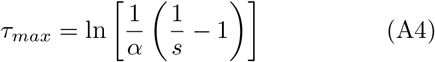

and the maximum number of daily cases

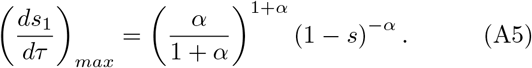

Taking into account the replacement of variables (3-5), this gives the point of the maximum *t_max_* and its peak value:

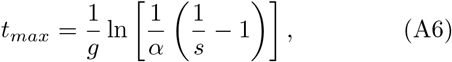

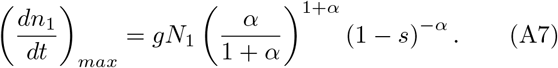

## Notes

### Competing Interest Statement

The authors have declared no competing interest.

### Funding Statement

No external funds was received.

### Author Declarations

No approvals are required

### Summary of Updates

Appendix B removed

